# Cardiovascular Disease in the Peruvian Highlands: Local Perceptions, Barriers, and Paths to Preventing Chronic Diseases in Andean Adults

**DOI:** 10.1101/2021.05.25.21257458

**Authors:** Giuliana Sanchez Samaniego, Stella Hartinger Peña, Paula Skye Tallman, Daniel Mäusezahl

## Abstract

**Objectives:** Public health interventions can be improved by understanding peoples’ explanatory models of disease. We explore awareness and perceptions of cardiovascular diseases (CVD) and options for preventive actions in young rural adults highlanders.

**Methods:** 46 men and women from communities in Cajamarca, Peru were purposively selected to participate in eight focus groups, where participants discussed their understanding and perceived causes of CVD as well as barriers and pathways to healthy lifestyles.

**Results:** Fresh foods, physical activity, unpleasant emotions, and healthcare access were cited as important determinants of healthy lifestyles. Barriers to healthy diets included lacking nutritional knowledge, fluctuating food prices, and limited access to foodstuffs. Women felt particularly vulnerable to CVD and identified gendered barriers to manage stress and engage in leisure sports. Low health literacy, distance and poor doctor-patient relationships prevented participants from fully accessing healthcare.

**Conclusions:** CVD prevention should consider local knowledge of these diseases and of healthy lifestyles and harness ongoing programmes that have successfully promoted good nutrition in children and pregnant women. In concert with public-private parterships, governments should include health prevention for the entire family.

## Introduction

Cardiovascular diseases (CVD) accounted for approximately 18.6 million deaths in 2019 and are considered the leading cause of death globally [1]. CVDs are a special challenge for low and middle-income countries, where they coexist with a high prevalence of infectious diseases and maternal health conditions [2], and burden health systems that are plagued by inequitable access to care [3].

Peru is a case in point. The country faces the double challenge of malnutrition and infectious diseases and increases in CVD [4]. The number of studies of CVD in Peru is growing [5-8] and prior research shows that highland populations have a lower CVD risk in comparison to low-altitude sites [9]. However, current research predominantly utilises an epidemiological perspective to understand who is at greatest risk and often misses individual perceptions of CVD and locally relevant factors that limit the adoption of healthy behaviours.

To address this gap, this study documents local understandings of CVD and healthy lifestyles among Andean adults in Cajamarca, Peru. We explore participants’ personal awareness of CVD and the barriers to adopting healthy behaviours using the Health Belief Model (HBM). The HMB framework focuses on understanding personal attitudes, beliefs and practices, which is crucial for the development of intervention approaches that are culturally acceptable, people-driven and converge people closer to the health system [10-12]. We see this as a first step to determining locally relevant behavioural programmes that can help slow the enormous wave of chronic disease risk that threatens the health of Andean adults.

## Methods

### Study Setting

This study was carried out in the provinces of San Marcos and Cajabamba in the Cajamarca region, in the northern highlands of Peru, 1800-3900 MASL [13]. These provinces include semi-urban and rural communities. The majority of the population lives in houses made of clay-sand walls and earthen floors. Farming and livestock are the principal source of income [13]. Several governmental programmes are active in the area, including the nutritional interventions “*Vaso de leche*” (for pregnant women and children under 6-years-old) and “*Qaliwarma*” (for school age children), and two conditional cash transfer programmes, “*Juntos*” (for pregnant women and families with children under 15 years old), and “*Pension 65*” (for adults over 65 years old that live in extreme poverty) [14]. Different ministries, including the Ministry of Development and Social Inclusion, the Ministry of Health and the Ministry of Economy and Finance, implement these programmes [14].

### Study Design and Participants

We conducted focus group discussions with adults in the Province of San Marcos and Cajabamba. Participants were parents of children enrolled in a previous study in the area [13]. Additionally, at least one parent had undergone a metabolic syndrome examination [15].

### Recruitment

We used purposive sampling to select participants. Subjects from eight out of 68 local communities were invited to participate.The field supervisor and a fieldworker visited the participants’ household and invited them to take part in the focus groups 3-4 days before the scheduled session. Between four to 10 male and female individuals participated in eight mixed focus groups [16].

### Focus Group Discussions (FG)

Focus groups were held at a central location such as a school or community centre, or at a participant’s house. During the focus group, the first author acted as the moderator, and two assistants worked as observers. The moderator facilitated the discussion and encouraged all participants to discuss the questions included in the FG discussion guide. The FG discussion guide was based on the six constructs of the Health Belief Model [10, 11]; perceived susceptibility, perceived severity, perceived benefits, perceived barriers, cues to action, and self-efficacy. We complemented this framework by adding three additional sections on patterns of distress, perceived causes, and gendered differences. The latter was added due to the higher prevalence of metabolic syndrome among women than men [15].

### Data Analysis

We recorded all FG discussions and two assistants took notes on verbal communication. Following each session, project staff transcribed the audio recordings, identified the participant speaking, and re-listened to the tapes to ensure accuracy. An inductive approach was used for data analysis. Data was organised using the Dedoose qualitative analysis software [17].

The codebook consisted of topics identified in the FG guideline and emergent themes from the data. The moderator and the two observers coded the data. If differences in coding appeared, consensus was reached after a team discussion. Similar responses between participants of the same or different focus group were identified from coded extracts. Extracts were organised into tables, with identifiers for the focus group, the participant number, and the theme code. Additional demographic and health information for the participants was available from the larger study this research was embedded in [13, 18].

### Ethics

The Universidad Peruana Cayetano Heredia (UPCH) Ethical Review Board approved the study.

Informed consent was obtained from all individuals included in the study. They all consented to the recording of the session.

## Results

### Characteristics of participants

A total of 46 participants (37 women and 9 men) partook in the eight FGs. Each FG lasted between 30 minutes and one hour. Demographic and health information are shown in Table 1.

**Table 1:** Demographic and health characteristics of young adult participants of the Provinces of San Marcos and Cajabamba, Peru 2017. Legend: SD: standard deviation MASL: metres above sea level

|  | Women |  | Men |  |
| --- | --- | --- | --- | --- |
| | N | Mean ± SD or N (%) | N | Mean ± SD or N (%) |
| Age (years) | 37 | 29.1 ± 6.0 | 9 | 29.3 ± 4.0 |
| Complete secondary | 34 | 9 (27%) | 6 | 4 (67%) |
| Work in the agriculture |  |  |  |  |
| sector | 34 | 23 (68%) | 6 | 6 (100%) |
| Housewife | 34 | 6 (18%) |  |  |
| Living over 2500 MASL | 37 | 15 (41%) | 9 | 6 (67%) |
| Body mass index | 34 |  | 6 |  |
| Underweight |  | 0 |  | 0 |
| Normal weight |  | 15 |  | 3 |
| Overweight |  | 13 |  | 3 |
| Obese |  | 6 |  | 0 |
| Metabolic syndrome | 34 | 10 (29%) | 6 | 3 (50%) |

### Main Findings

#### Understandings of CVD

In all FGs at least one participant knew a person who suffered from a CVD, but there was dissatisfaction with the knowledge that was available and accessible about CVD. One man in his mid-30s stated, “Yes, we have heard about these diseases, we even know heart transplants exist. All of that. But we don’t have information to prevent these diseases, nor what they do, nor what causes them”.

Following up on why there is a lack of information, a woman in her mid-20s expanded that she had not received information on CVDs from the health services, “Because the health staff in this health post only cares about the main reason of your visit. For example, child growth control. They just examine that and it’s done”. Another man in his mid-30s expressed a similar sentiment, “About nutrition, about other diseases, for example, tuberculosis, HIV, cancer. For all those… yes, there is prevention… different pamphlets, but very few about the heart”. High levels of illiteracy are an additional factor complicating understandings of CVD. As a woman in her mid-30s stated, “The one who knows how to read, can read. But the one who doesn’t know, only observes”.

While the participants expressed dissatisfaction with the level of information available for CVD and cited that they have not had in-depth discussions of CVD previously, they still expressed knowledge about the causes and symptoms of CVD. Specifically, they mentioned that symptoms of CVD included agitation, pain, weakness and strong heart beating; and that the ultimate consequences of CVD were heart attacks and death. In all focus groups the participants stated that anyone could develop a heart disease, no matter their weight or age. Additionally, participants identified a number of drivers of CVD, which fell into four general categories: nutrition, physical activity, emotions, and access to healthcare.

### Nutrition

Study participants expressed that CVD is associated with being overweight and “having fat around the heart”. Interestingly, a woman in her early 30s stated, “When we have fat in the heart too, and is not necessary that we are fat” and other community members clearly articulated that being overweight or underweight could be a determinant of CVD. Another woman in her mid-30s expanded, “If you are overweight or you are underweight, it is equally bad.” and that “Nobody is free … no one is free neither the fat nor the skinny. I believe that not only the fat person gets sick, furthermore the skinny one may be sicker”.

Participants not only acknowledged that both under and overnutrition were problematic, but also made direct connections between nutritional health and food. A woman in her late-20s confided, “Sometimes, we only eat to fill our belly, without realizing that it is not truly nourishing us”. Further, participants explained how structural factors limited their ability to obtain good nutrition. A woman in her early-30s stated that, “Sometimes we eat things that are bad for us and we know that they are going to be bad for us. But we eat out of necessity because there is no other thing to eat”.

The identification of structural barriers to adopting a nutritious diet were common and included not being able to access healthy foods, not having the economic resources to purchase them, and lacking knowledge about how to prepare healthy foods. These barriers varied across geography and occupation. More than one third of the participants were farmers, but reported that they did not regularly consume products from their crops. A man in is his mid-30s told us that, “50% [of all goods] you have to buy and another 50% is produced in the field”.

Participants living in the low altitude valley (1800-2200 MASL) reported that they can grow all types of food. However, community members in higher altitudes are limited in their crop variety and also had issues with access to purchasing fresh food products such as vegetables, fruits and animal protein. Barriers to obtaining these products included the cost to reach a market (which could be between 30min– 2 hours away), time spent getting there, and issues with food storage. As a woman in her late-20s pointed out, “Food cannot be stored for a long time, maybe 2 or 3 days because we do not have a refrigerator, a place where to keep the food”.

On the other hand, local *bodegas* provide constant access to non-perishable foods and drinks, which are cheaper. As a man in his mid 30s quipped, “Mostly you decide what food to buy according to the economy, right?” But even buying foods at the *bodegas* was complicated because food prices changed every week because of the many intermediaries involved. As a woman in her mid-30s explained, “You know… the intermediaries always have to take advantage, right? They live off that, so sometimes they sell a product at a certain price and the next week they change it. Then, in the local store, items are more expensive.” Local challenges to accessing, purchasing, and storing foods lead people to preferentially buy non-perishable foods such as rice, noodles, oil, and sugar, especially those participants living in distant communities.

### Physical Activity

Participants indicated that physical activity was beneficial for their wellbeing and recognised it as a preventive measure for developing CVD. They mentioned being highly active because of their daily agricultural and farming work, which they enjoyed and kept them busy. For example, a man in his late-20s stated that, “We get health benefits, and economic benefits…. to be useful (for field work)”. However, participants explained that recreational physical activities such as sports were often limited due to lack of time, religious and local beliefs, and gendered factors. A man in his mid-30s shed light on this saying, “The word of God [local church] has forbidden us to play football. We always bet sometimes, right? Sometimes we have anger at the partner because he is beating us. Sometimes we do not play well.”

Women appear to face dual pressures. On the one hand, participation in sport appears to be expected of mothers. A mother in her late-20s expanded saying, “Sometimes in the schools, in kindergartens, the moms have to participate. There is no way you do not participate. If the babies see that their mother does not want to, then they will not want too. So, you have to participate, no matter what.” Yet there is also gendered pressure to refrain from sports as a woman in her mid-30s stated that sometimes woman are expected to stay at home and take care of their family and not spend leisure time in sports. Gendered dynamics in CVD risk factors were also evident in group discussions regarding the perceived role of emotional stress in CVD.

### Negative Emotions

Participants articulated that negative emotions could contribute to heart problems and also that knowing they have health problems can produce negative emotions. Speaking about a family member, a woman in her late-20s stated that, “My aunt… sometimes she suffers attacks. She has been told to avoid being melancholic, to avoid worries. Nothing, nothing. No worries, because if she is worried or melancholy she can die.” Indeed, participants connected both physical and emotional stress to strain on the heart. A young woman explained that, “Sometimes when you lose your patience. You feel that your heart beats faster at times like this. Sometimes you have anger, let’s say, it’s a moment …”, while a woman in her late 30s expressed a similar sentiment in relation to physical activity, saying, “When you walk and feel agitated, then you feel pain and sometimes “stitches”. Well then, you think it is the heart, right?”.

These quotes indicate that negative emotions or over-exertion create heart problems, but participants also indicated that simply knowing about having heart problems can produce substantial household distress. Indeed, a number of participants expressed that they would feel sad and worried if they were diagnosed with any CVD as the costs of treatment, or losses due to death, could drive economic insecurity for the entire household. Women worried about themselves and their partners saying, “If my husband gets sick, what should I do? Who works? Who maintains us?” and “If my husband gets sick, I have to spend money, I do worry.”

Discussions in the FGs revealed why women appeared to be more prone to this type of worrying. A woman in her late-20s explained, “It seems to me, I say it is because men go to the farm, they occupy themselves there, they forget about everything. On the other hand, the women stay at home and sometimes we have worries about the children… everything, that can also be it [a risk factor of CVD] or maybe not”. A man in his late-30s expressed a similar explanation, musing that “Maybe… [Women are at higher risk] because sometimes we, men, go to work, it is a single job and the ladies stay at home, they work different tasks…they worry sometimes and think “There are things that I don’t have…. I do not have money”.

Finally, participants expressed that they would rather not know about a potential cardiovascular condition because of the stress it might cause, saying, “It is better to not know (that you have a CVD). Just stay quite during the pain, then it goes away and you forget…. if you know what you have, it is worse.” However, even for those who are interested in knowing about their CVD risk, there are substantial barriers to accessing knowledge, affordable treatment, and care at the local health posts.

### Healthcare

Similarly, to the challenges reported in accessing healthy fresh foods, participants cited costs associated with transport and food as a barrier to healthcare. As a result, participants stated that they typically only went to the clinic for a pressing health issue and not for regular check-ups. A woman in her late-20s stated, *“*If it does not hurt, I don’t go to the doctor. If it hurts, I will go to check what is going on. On the contrary, we should have general check-ups, for example annually.” Appointments at the health post also needed to be scheduled in person, potentially creating two lengthy and costly trips, and creating a high initial barrier to seek care.

In addition to challenges in accessing care, participants criticised the quality of patient-provider relationships. They felt that they were not treated well nor receiving proper attention because health professional were inexperienced and prescribed painkillers for every condition. According to a woman in her mid-20s, previously, “There was a good nurse, but they relocated her to the hospital. And here, we only have trainees. This is what happens here at the health post”. and when she does visit the health post, “Oh, this hurt, oh, paracetamol, ibuprofen. All the time it is the same medicines we receive. When we go with our children, we already know what they will prescribe to us.”

More problematically, participants were receiving conflicting medical advice that was not evidence-based. A woman in her mid-30s revealed that

> “I went to the health post with my results [lipids and glucose measurements]. They [health professional] told me: “No, there is no medicine, you can only take warm water before eating”. I told them I couldn’t eat in the morning. They told me that anyway I should eat a little bit…. but I should eat. They told me to start eating less but not to stop eating. They told me better to buy wine and drink a glass before going to sleep to get rid of the fat. But, I have not seen any results.”

This type of medical advice appears to be intersecting with local explanatory models for the treatment of CVD. One woman in her early 30s stated,”For example, if the heart is not well and they (health professional) tell you it is because of the fat.. then you try to eat things that do not affect your heart. Right? I know that you have to drink pineapple juice and because you know this is good for the heart then you drink it…, this is a medicine…. right? A similar sentiment was expressed by a woman in her mid-20s, “Yes, sometimes one feels bad. You can try to look for flowers, and drink flower waters, you can use also lemon, orange, custard apple”

It is unclear whether these remedies were coming from healthcare practitioners, traditional healers, or fellow community members. What is apparent is that the public health messaging for preventing and treating CVD is not completely clear. Despite this, participants related CVD to nutrition and exercise and many participants showed an interest in adopting healthier diets and increasing physical exercise. However, they also identified a number of substantial challenges to enacting these behaviours in the short and long-term, which are key issues to address in launching successful behavioural interventions to prevent CVD in Andean communities in Peru.

## Discussion

In this paper, we explored how Andean adults in Peru understood CVD, including the drivers of CVD risk, and the barriers to adopting healthy lifestyles that can reduce risk. While the understandings of CVD were basic, study participants stated that nutrition, physical activity, negative emotions, and healthcare were important drivers of CVD risk. Here we expand on these findings with particular attention to how they can inform the National Guideline of Prevention and Control of Non-Communicable Diseases and their strategies for vulnerable high-altitude populations [19].

Many of the participants in this study knew that CVD was potentially life-threatening and expressed a desire to be better informed. However, participants made it clear that health communication strategies via text or print-outs would not reach illiterate community members. Instead, they suggested that nutritional workshops with demonstrations of healthy cooking and nutritional discussion groups could effectively engage community members.

Such nutritional workshops, if designed holistically, could address many of the barriers to the adoption of healthy eating behaviours [20, 21]. Participants understood that healthy diets mean eating various types of food in proper amounts but felt their ability to adopt these diets was limited because they did not know how to prepare healthy meals and could not afford and access readily diverse food items. While there are existing programmes providing food to children and pregnant women in these areas, including *Vaso de Leche* and *Qaliwarma*, adults without children are notably excluded. As main programme targets, women have to comply with regular maternal and children health checkups and ensure children’s school attendance [14], making them central stakeholders for such programmes and a regular point of contact with the health system. Women are the key health information channels into the family. As primary knowledge-bearers and with their expressed desire for nutritional skill-building they are the ideal partners for people-centred engagement in CVD prevention in remote Andean communities.

We suggest that additional studies are conducted in close collaboration with Andean women to design nutritional interventions that can improve health and wellbeing at all life stages and are appropriate for this unique context. There are existing governemntal programmes such as Haku Wiñay (“My enterprising farm” in Quechua language), which support the raising and trading of domestic animals and entrepreneurship in small-scale agriculture[14]. Is it possible to expand these programmes to support women in small-scale subsistence activities that can provide healthy foods that can dually reduce CVD risk in adults and undernutrition in children? Identifying nutrient-rich and culturally-valued crops, suitable for both, home consumption and trading can enhance food security and strengthen income generating capacities. Additionally, opportunities exist to build on public-private partnerships for CVD prevention. For example, the national dairy company collects milk from the remotest areas of the Andes at least weekly. Considering that the company maintains this regular supply chain with remote outreach, building public-private partnership could help providing healthy foods to highlanders who otherwise travel only monthly or bi-weekly to local markets

However, to be feasible, such initiatives need to be linked to system-wide changes and interventions. Peru has successfully executed multi-level actions to address childhood undernutrition and anemia using a combination of programmes in the areas of health, education, cash transfer, water and sanitation, housing, and agriculture [22, 23]. CVD prevention action would need the same approach and could piggy back off of ongoing programmes with already established contacts within the health system. Specifically, CVD risk needs to be assessed at the peripheral primary care level measuring by anthropometrics and blood pressure, and short risk questionnaires [24, 25]. Health data should then be saved in a national repository of chronic conditions similar to the “Wawared” system for pregnant women and children[26]. With surveillance systems in place, monitoring, evaluation and access to treatment can become feasible in distant populations. Task-shifting [27] is another option that has contributed to maternal and child health programmes in Peru in the past and could be utilised for CVD interventions. Task-shifting entails working with community health workers (CHW) to bring preventative strategies to the periphery and is potentially helpful in settings with health workforce shortages [28]. Task-shifting requires the training of CHW in new skills, the use of standardised protocols, and the provision of adequate equipment for CVD prevention and communication. In this way, health professionals can expand from a child-centred health approach to include adult health. The above recommendations are inherently people-centred and community-based, however they are the first approach intro creating more comprehensive national guidelines for CVD prevention [19].

In 2015, non-communicable diseases were included in a global development agenda [8]. This target requires multi-level action beyond the health sector, as showed by the successf of child nutrition interventions in Peru [22, 23].Thus, system-wide health interventions are possible in the country and can dually address CVD risk in adults and continue supporting the nutritional health of children. Such approaches designed in partnership with local women, could also potentially address emotional distress as a notable locally perceived CVD risk factor, by promoting health and wellbeing and improving food security and access to care.

The study was conducted in young adults in their 30’s and perceptions of CVD may differ from older individuals. Villagers participating in the FGs knew each other, potentially influencing their participation. However, they collectively reflected a community view, which was particularly valuable when discussing locally feasible and acceptable intervention approaches.

Most of the FG participants were women but as household managers and family-health knowledge-bearers with the most regular contact with health facilities we believe they and their participating husbands closely represent the broader community perspective on NCD health matters.

This research revealed that there are many feasible entry points of intervention that can prevent Andean community members from succumbing to the wave of CVD risk that is currently engulfing the globe. Specifically, local women are key stakeholders who can influence their own health and the health of all family members. Therefore, they must be empowered with the knowledge and resources to enact healthy lifestyles. We see substantial opportunities for this to occur by expanding existing government programmess to make healthy foods and healthcare accessible, promoting healthy lifestyles and by building on public-private partnerships that can support access healthy food for all community members.

## Data Availability

The data generated and/or analysed during the current study are not publicly available but are available from the corresponding author on reasonable request.

## Acknowledgements

We thank the study participants for their valuable time and kind participation. We appreciate and thank the local authorities of the RED IV de Salud San Marcos, for their continuous support. We also express our gratitude to Mrs Angelica Fernandez for helping and organising community meetings and Karen Meza and Forlly Chavez for supporting the coding of the transcripts and organization of extracts

